# Identification of full-length circular RNAs linked with therapy resistance of pediatric cancers

**DOI:** 10.64898/2025.11.28.25340833

**Authors:** Chloé Bessiére, Loélia Babin, Elissa Andraos, Johannes Markus Riepl, Annabell Szymansky, Marco Lodrini, Hedwig E. Deubzer, Angelika Eggert, Cyril Quivoron, Charlotte Rigaud, Véronique Vérgé, Stéphane Pyronnet, Laurence Lamant, Fabienne Meggetto, Christine Gaspin, Steffen Fuchs

**Affiliations:** Inserm, UMR1037 CRCT, Toulouse, 31100, France; Université de Toulouse-Paul Sabatier, UMR1037 CRCT, UMR5071 CNRS, Toulouse, 31100, France; Equipe labelisée FRM, Toulouse, France; Department of Pediatric Oncology and Hematology, Charité - Universitätsmedizin Berlin, corporate member of Freie Universität Berlin and Humboldt-Universität Berlin, Berlin, 13353, Germany; German Cancer Consortium (DKTK), partner site Berlin, a partnership between DKFZ and Charité-Universitätsmedizin Berlin, Berlin, 13353, Germany; Berlin Institute of Health at Charité – Universitätsmedizin Berlin, Berlin, 10178, Germany; Experimental and Clinical Research Center (ECRC) of Charité and Max-Delbrück-Center of Molecular Medicine in the Helmholtz Association, Berlin, 13125, Germany; Max-Delbrück Center of Molecular Medicine in the Helmholtz Association, Berlin, 610101, Germany; University Hospital Essen, University of Duisburg-Essen, Essen, 45147, Germany; Translational Research Hematological Laboratory, AMMICA, INSERM US23/CNRS UMS3655, Gustave Roussy Cancer Campus, Villejuif, 94800, France; Department of Pediatric and Adolescent Oncology, Gustave Roussy Cancer Institute, Paris Saclay University, Villejuif, 94800, France; Department of Medical Biology Pathology, Gustave Roussy Cancer Campus, Villejuif, 94800, France; Univ Toulouse, INRAE, BioinfOmics, GenoToul Bioinformatics Facility, Castanet-Tolosan, 31326, France; Univ Toulouse, INRAE, UR 875 MIAT, Castanet-Tolosan, 31326, France

**Keywords:** ALK, non coding RNA, lymphoma, neuroblastoma, circRNA, pediatric oncology, Nanopore sequencing

## Abstract

Resistance to cancer treatment remains the leading cause of cancer-related deaths. In tumors with low mutational burden such as pediatric cancers, alternative transcripts, including circular RNAs (circRNAs), have been identified as involved in treatment resistance. However, their isoforms are often missed by commonly used short-read sequencing. Here, we employ long-read sequencing to identify full-length circRNA isoforms associated with resistance in *ALK* -driven pediatric cancers. Using cell models and a cohort of *ALK* -translocated anaplastic large-cell lymphoma (ALK+ ALCL) patients, two circRNAs were detected as specifically upregulated in resistant cases and associated with worse disease outcomes. Similar findings were observed in the pediatric cancer neuroblastoma. These circRNAs were also more abundant in liquid biopsies from ALKi-resistant ALK+ ALCL and neuroblastoma patients. This demonstrates that long-read sequencing allows for uncovering disease-relevant circRNA isoforms that could serve as biomarkers for resistance detection in a clinical setting.

## Introduction

Resistance to cancer treatment still limits patient survival, despite new treatment strategies, including targeted therapies [1]. While many resistance mechanisms based on genetic mutations have been identified in adult cancers, our understanding of those underlying for pediatric cancers is still limited due to a relatively low mutational burden in comparison to adult cancers [2], such as in *ALK* -translocated anaplastic large-cell lymphoma (ALK+ ALCL) and neuroblastoma.

ALK+ ALCL is a rare form of non-Hodgkin’s lymphoma that mainly affects children or adolescents. It is generally characterized by a translocation involving the anaplastic lymphoma kinase *ALK*, a surface tyrosine kinase receptor, which activates oncogenic signaling pathways. In cases where first-line chemotherapy is ineffective, ALK inhibitors (ALKi) are now commonly used as second line targeted treatment. However, only a limited number of studies have investigated emerging resistance to ALKi treatment [3]. Few mutations of *ALK* or genes of downstream signaling pathways have been identified as origin of resistance mechanisms to ALKi [4], which however do not describe all resistant cases [5]. Neuroblastoma is a rare solid embryonal cancer arising in young children that shows *ALK* aberrations in 10-15% of cases [6]. Alterations of *ALK* occur as amplifications or mutations and can be targeted with ALKi similarly to ALK+ ALCL [7]. Predictive biomarkers for the detection of ALKi resistance and monitoring of the ALKi therapy are currently missing.

While genomic mutations are rare in pediatric cancers in comparison to adult tumors, noncoding RNAs, such as circular RNAs (circRNAs) have recently been identified by us and others to be associated with neuroblastoma cell survival, proliferation [8] and chemotherapy resistance [9]. CircRNAs are noncoding RNA circles that are generated by a form of alternative splicing (AS) termed back-splicing that covalently links a 5’ downstream end and a 3’ upstream end to form a back-splice junction (BSJ) that leaves no free end [10]. They are important for gene expression regulation, show increased stability against degradation due to their closed covalent ring-structure and are detectable in cancer tissues and body fluids [11]. The BSJ is the only feature that distinguishes a circRNA sequence from the sequence of a linear RNA and is used to identify chimeric reads containing the BSJ of the circRNA. Although most known circRNAs were discovered using short-read sequencing, this strategy does not reveal the full-length sequence of circRNAs, AS events and the different isoforms that are needed to fully understand the function of a specific circRNA. Recent studies reported long-read sequencing strategies by using the Oxford Nanopore platform (ONT) to detect full-length circRNAs, which revealed the existence of many isoforms and specific AS events of circRNAs [12–15].

Here, we aimed to identify circRNA isoforms associated with the emergence of therapy resistance in *ALK* -driven pediatric cancers. As model diseases, we used ALK+ ALCL and neuroblastoma. By sequencing full-length circRNAs [16], we analyzed a panel of ALK+ ALCL cell lines resistant and sensitive to ALKi. By this means, we detected 9,702 circRNAs, of which two were associated with ALKi resistance. Our results show that the two circRNA candidates were associated with a poor patient outcome and were more abundant in liquid biopsies of patients resistant to ALKi. Similar findings were obtained in neuroblastoma. Ultimately, this research highlights the use of long-read sequencing approaches to detect predictive circRNA biomarkers in a clinical context.

## Results

### A higher number of circRNA isoforms was identified in ALK+ ALCL cell models resistant to ALK inhibition

To identify circRNA isoforms associated with resistance to ALKi in ALK+ ALCL, we employed cell line models sensitive or resistant to treatment with the ALKi crizotinib that is used in clinical routine for ALK+ ALCL patients. Crizotinib-resistant Karpas-299 and SUP-M2 cells were generated by culturing them with sublethal concentrations of crizotinib. Employing the same approach, we obtained resistant SU-DHL-1 and COST cell models.

Using our previously adapted protocol to enrich for circRNAs and sequence them in full-length with the Oxford Nanopore long-read sequencing platform (see methods) [16], we analyzed 4 pairs of ALK+ ALCL sensitive and resistant cell models **(Fig. 1a)**. The enrichment of circRNAs in the samples was confirmed by quantitative RT-PCR (qRT-PCR) targeting an abundant circRNA and control transcripts **(Supplementary Fig. 1a,b)**. The number of sequenced reads (median of 1.86 and 1.94M for sensitive and resistant groups respectively), and read length were not significantly different between the sensitive and resistant groups **(Supplementary Fig. 1c,d)**, which shows the comparability of the samples. To minimize false positive circRNA isoform candidates, we employed two different circRNA detection tools, circFL-seq [14] and CIRI-long [15]. In total, we could detect 9,702 full length circRNAs that were supported by both tools (30,532 circRNAs predicted by circFL-seq and 25,496 by CIRI-long; **Fig. 1b), Supplementary Data 3 and 4**. Interestingly, we detected in general more circRNAs in resistant cell models **(Supplementary Fig. 1e,f)**, and they tended to be longer (mean length in sensitive: 470.8 nt, vs. resistant: 508.3 nt, **Fig. 1c)** and more diverse, with more different isoforms, when comparing with sensitive models (total different isoforms of 4,898 and 6,775 in sensitive and resistant models, respectively, **Fig. 1d)**. Comparisons with the public database circAtlas [17] showed that we detected 4,902 not previously annotated circRNAs (50.6% of all common detected circRNAs, in line with previous reports [18]).

**Fig. 1.**
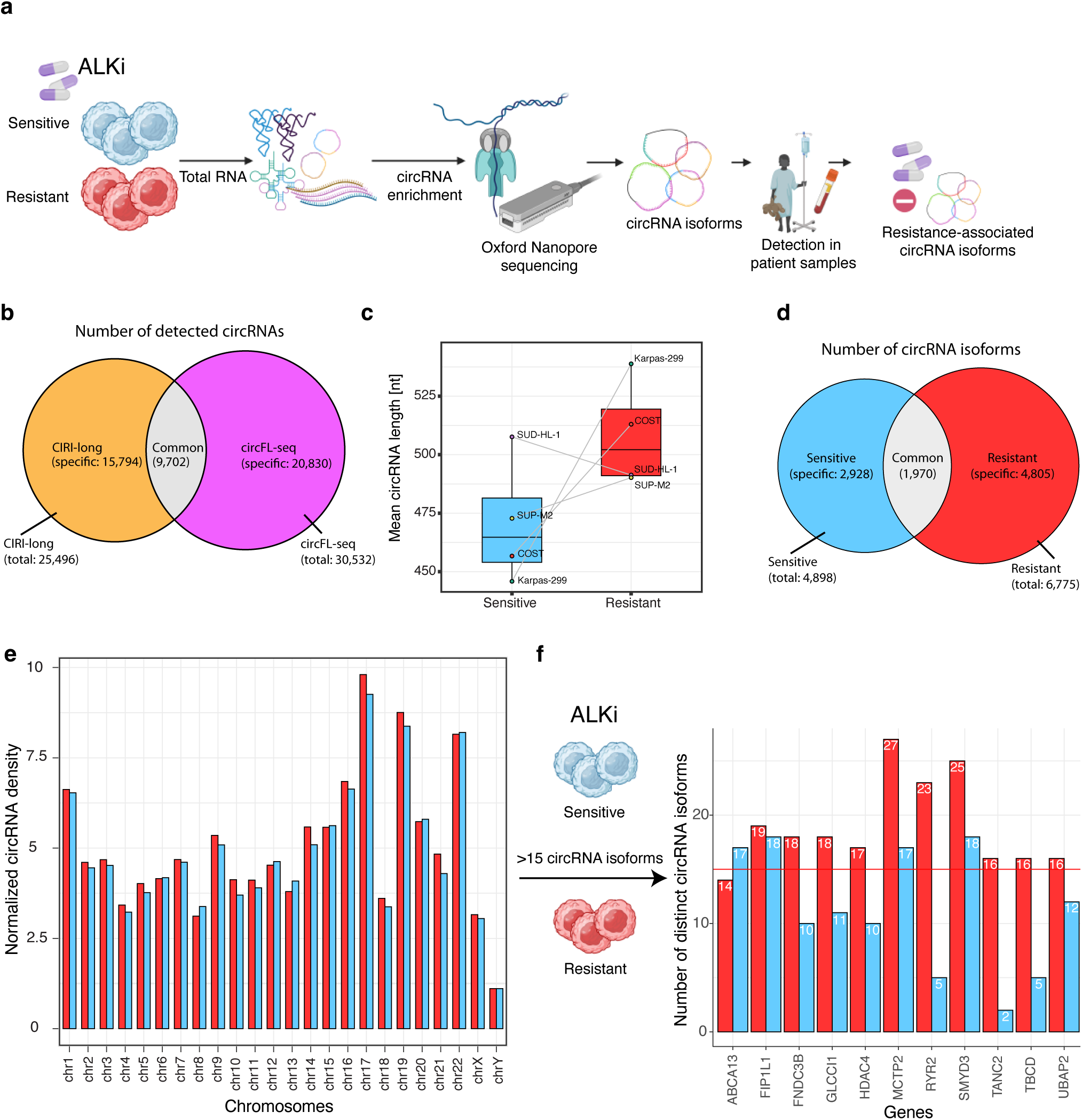
circRNAs show numerous alternative splicing events associated with therapy resistance. **a,** The scheme shows the workflow of the study. Enriched circRNA fractions of 4 pairs of sensitive and resistant ALK+ ALCL cell models (Karpas-299, SUP-M2, SU-DHL-1, COST) were sequenced in full-length using ONT. Identified resistance-associated isoforms were validated *in vitro* and *in vivo*. **b,** circRNA isoforms were bioinformatically detected with the tools CIRI-long and circFL-seq in the ALKi sensitive (n=4 biologically independent samples) and resistant (n=4 biologically independent samples) groups. The overlap of detected circRNAs is shown as a Venn diagram. circRNAs commonly detected are used as a robust dataset for downstr_2_e_1_am analysis. **c,** Mean length (nt) of circRNA isoforms by cell line and in the ALKi sensitive (blue) and resistant (red) groups. Data are presented as boxplots. **d,** Total number of distinct circRNA isoforms in the ALKi sensitive (blue) and resistant (red) groups. The overlap of circRNAs isoforms between the 2 groups is shown as a Venn diagram. **e,** Density of total distinct circRNA isoforms per chromosome was calculated and normalized to the total coding gene length for each chromosome. The normalized density for both sensitive (blue) and resistant (red) cell models per chromosome is reported. **f,** Hotspot genes that produce more than 15 distinct circRNA isoforms are shown in the 2 groups from e. In boxplots the center line represents the median, boxes indicate the interquartile range, the whiskers show the 1.5 interquartile range. Data are represented as a barplot showing the absolute number. Source data are provided as a source data file.

Motivated by our previous study in biopsies of the pediatric cancer neuroblastoma [8], showing that several genes and chromosomal regions are hotspots for the generation of circRNAs, we wondered whether the same is the case in our ALK+ ALCL dataset. Several hotspot chromosomes, such as chromosomes 17, 19 and 22, were identified for both sensitive and resistant cell models **(Fig. 1e)**. While most annotated genes produced only a few circRNA isoforms, as previously described [8], we detected 11 genes that produced more than 15 different circRNA isoforms either in sensitive, or resistant cell models. We identified 10 of those circRNA hotspot genes with more isoforms in resistant cells and only 1 in sensitive ones **(Fig. 1f)**. For instance, the *RYR2* gene, located on chromosome 1, produces 23 different circRNAs in our samples. We detected *RYR2* as a hotspot gene with a higher number of circRNA isoforms specifically in resistant cells and with only 5 isoforms in sensitive cells. Taken together, our results indicate that ALK+ ALCL cell models resistant to ALKi express a higher number of more diverse circRNAs.

### Alternative splicing events of circRNAs are more frequent in resistant ALK+ ALCL cell models

We hypothesized that internal AS events in circRNAs may be associated with resistance and investigated this by leveraging our long-read sequencing data, using the common circRNAs predicted by both tools for higher confidence. Hereby, we focused on the most common AS events: complete intron retention, exon skipping and use of novel (non-annotated in the reference transcriptome) exons **(Fig. 2a)**. For all 3 classes of events, we compared resistant with sensitive models. In general, we detected a higher number of the different AS events in the ALKi resistant group **(Fig. 2b, Supplementary Fig. 2a, 3a)**. For example, we found 55 circRNAs with a fully retained intron event. Of those events, 42 were only found in resistant cell models (11 in the sensitive group, 2 in both groups; **Fig. 2b)**. Then, we selected one highly expressed event as an example of each class of AS: circELMOD3 (7,8,9,RI,10), with a full retained intron (RI) event between exon 9 and 10, circARNTL2 (2,3,4,NE,5) with a novel exon (NE) between exon 4 and 5 and circZNF207(7,8,10), in which exon 9 is skipped. These circRNA variants are annotated as isoforms in circAtlas **(Fig. 2c, Supplementary Fig. 2b, 3b)**. Since no specific database allows for a large scale systematic comparison of circRNA AS events with their exact genomic coordinates, we also queried all the annotated splicing events from the VastDB atlas [19]. This revealed that more than 150 of our circRNA AS events were present in VastDB, suggesting that this database contains AS events derived from linear as well as from circular RNAs.

**Fig. 2.**
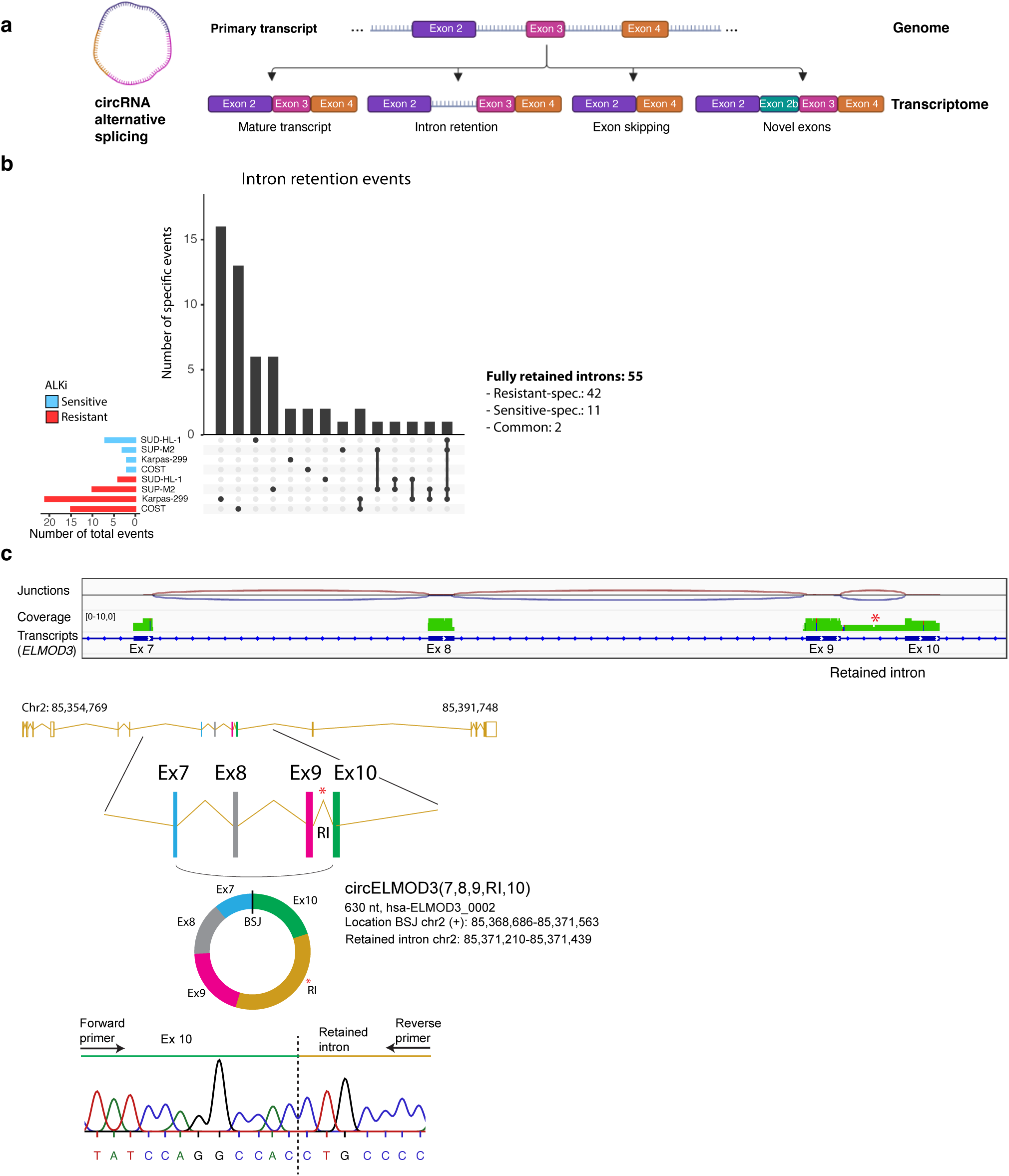
circRNAs show numerous alternative splicing events associated with therapy resistance. **a,** The scheme shows the alternative splicing e2v2ents that are considered for the analysis: intron retention, exon skipping and novel exons events. **b,** The upset plot shows the total number of intron retention events by group of cell models (blue, ALKi sensitive and red, ALKi resistant, each n=4 biologically independent samples) and the events specific or common (indicated by a connecting line) to the different cell models analyzed by long-read sequencing. **c,** The genome browser view shows exemplified circELMOD3(7,8,9,RI,10) with an intron retention event and the validation by Sanger sequencing of the junction between exon 10 and intron 9 in Karpas-299 cells. Details on the circRNA ID (circAtlas 3.0), length (nt) and coordinates are also given. Source data are provided as a source data file.

All three selected events were validated by PCR and Sanger sequencing **(Supplementary Fig. 3c)**. Two of the three AS events in circRNAs were stable after an exonuclease treatment, which indicates that they are derived from a circRNA, while linear controls were degraded. Summarizing, these results indicate that resistant ALK+ ALCL models show more AS events that increase the diversity of circRNAs in resistant cell models.

**Fig. 3.**
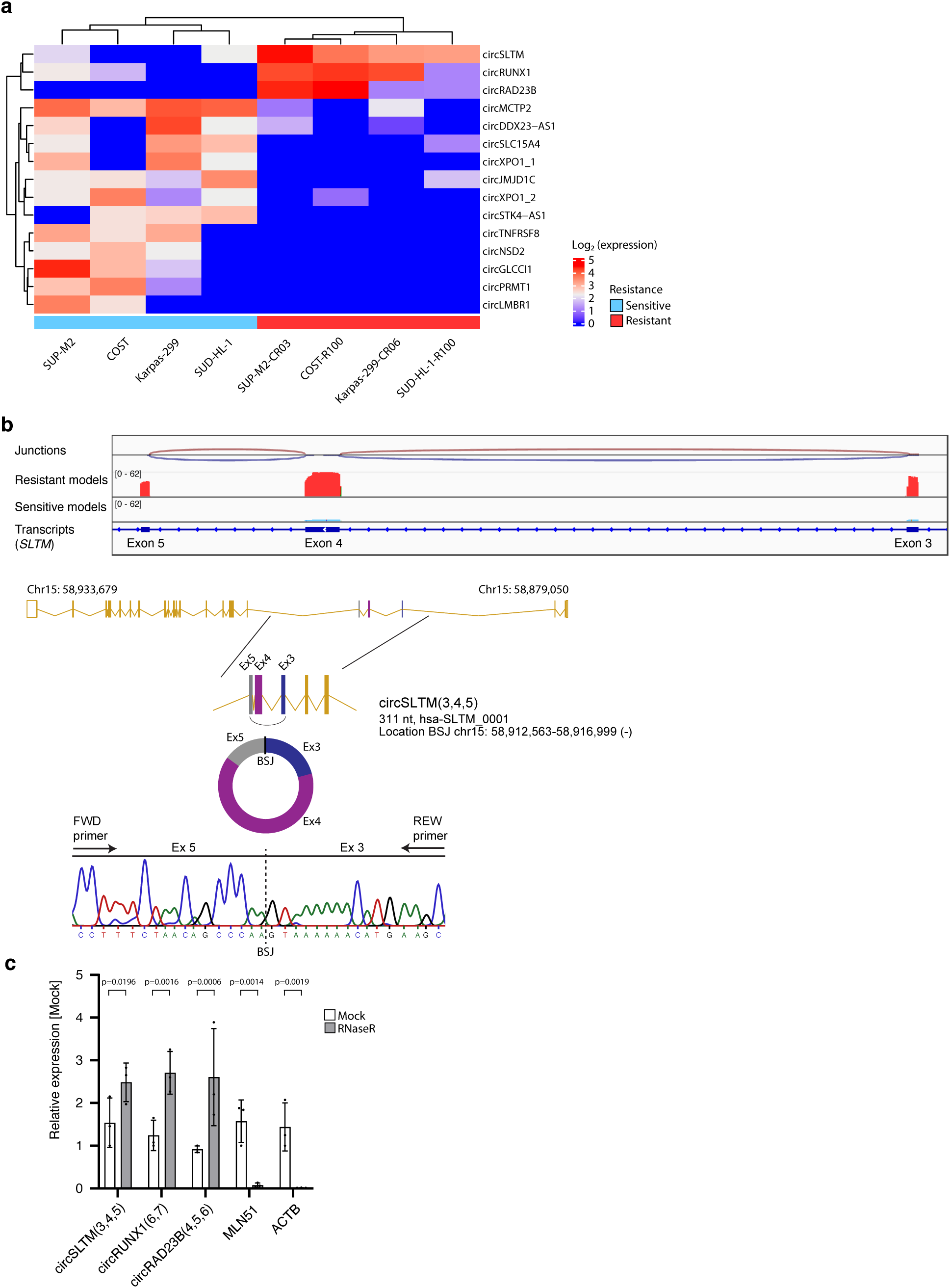
Nanopore sequencing identifies circRNA isoforms in full-length that are more diverse in ALKi resistant ALK+ ALCL cell models. **a,** Heatmap of the differentially expressed circRNA isoforms identified by Nanopore sequencing in ALKi sensitive (n=4 biologically independent samples, blue) and resistant (n=4 biologically independent samples, red) cell models. Only significantly different circRNAs are considered (p *<* 0.05). **b,** The genome browser view shows circSLTM(3,4,5) and its expression in resistant (red) and sensitive (blue) cell models. The validation of the back-splice junction (BSJ) by Sanger sequencing in Karpas-299 cells is shown. **c,** Validation of circularity of circRNA candidates by treatment with the exonuclease RNaseR in Karpas-299 cells and subsequent qRT-PCR in comparison to linear controls (n=3 biologically independent experiments, Data are presented as mean ± SD, Two-way ANOVA test). Source data are provided as a source data file.

### CircRNA isoforms are specifically upregulated in ALK+ ALCL cell models resistant to ALK inhibition

We aimed to find circRNA isoforms that were specifically expressed in resistant cell models and might serve as biomarkers associated with resistance to ALKi. Of a total of 2,651 circRNA candidates included in the analysis, we detected 15 full-length circRNA isoforms differentially expressed **(Fig. 3a)**. Three of the exonic circRNAs were specifically upregulated in the resistant cells: circSLTM(3,4,5), circRUNX1(6,7) and circRAD23B(4,5,6). Of those, circSLTM(3,4,5) showed the highest and most consistent expression among all 4 resistant cell models. The 3 circRNAs are identified in circAtlas and were validated *in vitro* by qRT-PCR and Sanger sequencing of the back-splice junction in Karpas-299 cells **(Fig. 3b, Supplementary Fig. 4a,b,c)**. Exonuclease treatment validated the circular character of the circRNAs **(Fig. 3c)**. Analysis of short-read sequencing data allowed for a global quantification including linear RNAs of the 4 cell line pairs that we created. No significant expression difference of SLTM, RUNX1 and RAD23B mRNAs was detected between the sensitive and resistant groups **(Supplementary Fig. 4d)**. This suggests that not the general expression of the genetic locus, but the circRNAs are specifically upregulated in resistant cell models. In total, these results show that 3 circRNA isoforms are specifically higher expressed in ALKi resistant ALK+ ALCL cell models.

### Resistance-associated circRNAs are linked with an unfavorable outcome of patients with ALK+ ALCL

Prompted by the higher expression of the 3 circRNAs in resistant cell models, we hypothesized that a similar expression pattern would be observed in clinical samples of patients with ALK+ ALCL. Since circRNAs show increased stability in body fluids, we aimed to detect them in liquid biopsies of ALK+ ALCL patients. Peripheral blood samples from a small group of ALK+ ALCL patients that showed resistance to ALKi were compared to blood samples from patients at diagnosis and from healthy donors as controls. This revealed a higher abundance of circSLTM (3,4,5) and circRUNX1 (6,7) in cell-free RNA of patients with resistance to ALKi **(Fig. 4a)**. Of note, circRAD23B (4,5,6) was not detected, which is why we focused on the 2 other circRNAs for the further analyses.

**Fig. 4.**
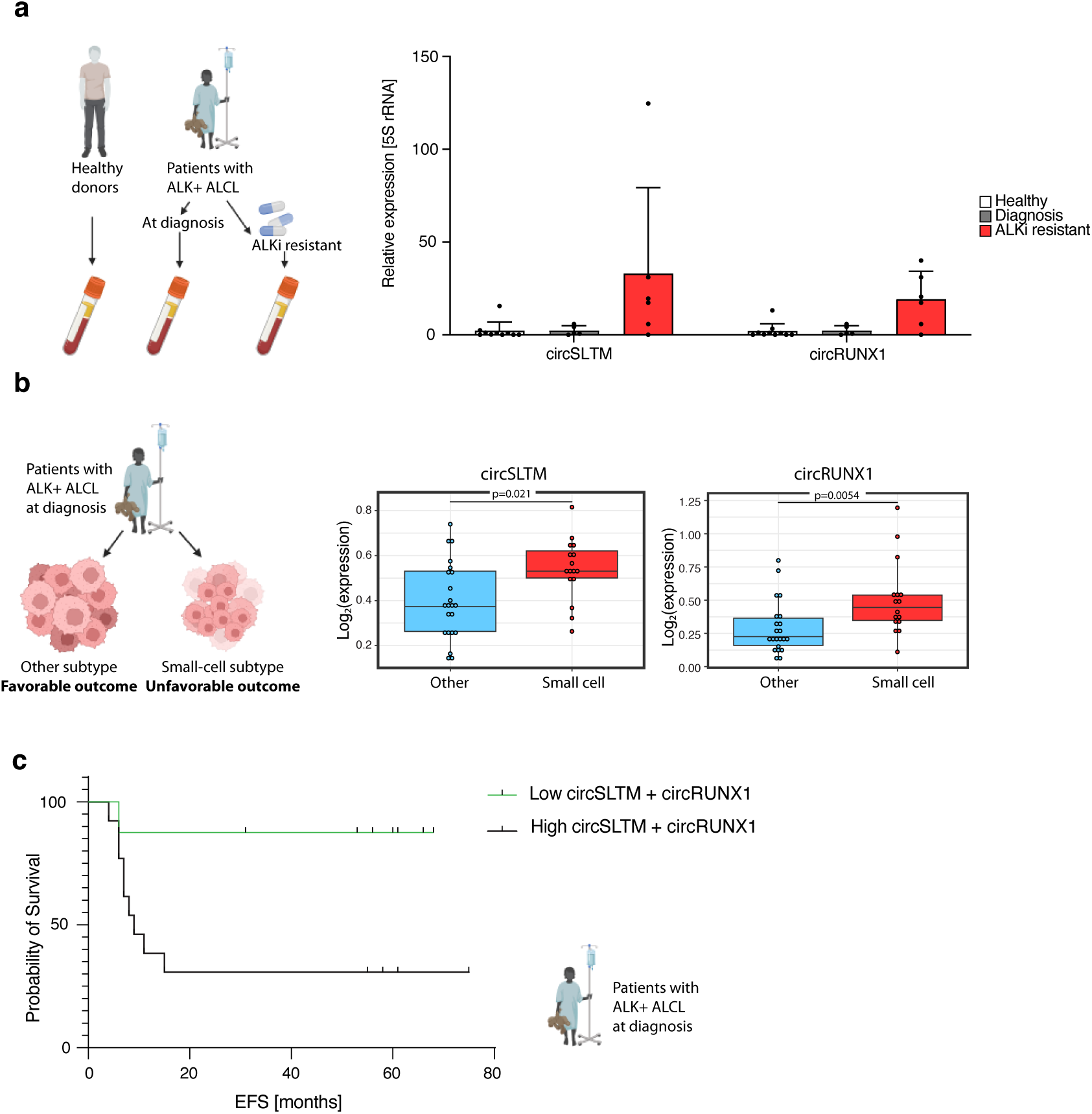
Resistance-associated circRNAs show higher abundance in clinical samples of patients with ALK+ ALCL. **a,** Serum samples of healthy donors (n=10 biologically independent samples) and from patients with ALK+ ALCL (n=4 patients) at diagnosis and after ALKi treatment showing a bad response were analyzed by qRT-PCR. The expression of circSLTM (3,4,5) and circRUNX1 (6,7) are shown. Data are presented as mean ±SD. **b,** Primary biopsies of patients with ALK+ ALCL with a small-cell containing (n=16 biologically independent samples) or other histological subtype (n=22 biologically independent samples) were analyzed by RNA-seq. **c,** The patients of b were divided in a group with both circSLTM (3,4,5) and circRUNX1 (6,7) low (n=8) vs. high (n=13) expression and the probability of event-free survival (EFS) in months is shown as a Kaplan-Meier curve. In boxplots the center line represents the median, boxes indicate the interquartile range, the whiskers show the 1.5 interquartile range. Source data are provided as a source data file.

We then re-analyzed total RNA short-read sequencing data of a cohort of tumor samples from 38 patients with ALK+ ALCL at diagnosis that we have previously generated [20]. The patients were separated based on the histological subtype of the lymphoma as evaluated by a trained pathologist. We distinguished a group containing small-cell components, which is a routinely assessed histological feature associated with an unfavorable prognosis [21], and a group that contains other subtypes (mainly large cell components) not associated with a negative outcome. Using our previously established kmer-based analysis approach to quantify circRNAs [22], we designed 40-mers specific to the BSJ of the two circRNAs. We detected circSLTM (3,4,5) and circRUNX1 (6,7) upregulated in tumors that contained small-cell components **(Fig. 4b)**. Further, a combined high expression of both circSLTM (3,4,5) and circRUNX1 (6,7) was associated with a shorter event-free survival (EFS, **Fig. 4c)**. Summarizing, these results show that the resistance-associated circRNAs were upregulated in clinical samples from patients with resistant ALK+ ALCL and associated with an unfavorable prognosis.

### A higher abundance of circRNA variants is detected in patients with ALKi-resistant neuroblastoma

We then explored whether circSLTM (3,4,5) and circRUNX1 (6,7) could be as well detected in the pediatric cancer neuroblastoma that recurrently shows *ALK* aberrations and can be treated with ALKi in these cases. Similarly to ALK+ ALCL, we collected peripheral blood samples of a small exploratory cohort of patients with neuroblastoma to test whether the circRNAs are detectable in liquid biopsies. Samples were obtained after treatment with ALKi and cell-free RNA was isolated. Both circSLTM (2,3,4) and circRUNX1 (6,7) were found more abundant in patients resistant to ALKi when compared to sensitive patients **(Fig. 5a)**. Again, circSLTM (2,3,4) showed higher levels than circRUNX1 (6,7).

**Fig. 5.**
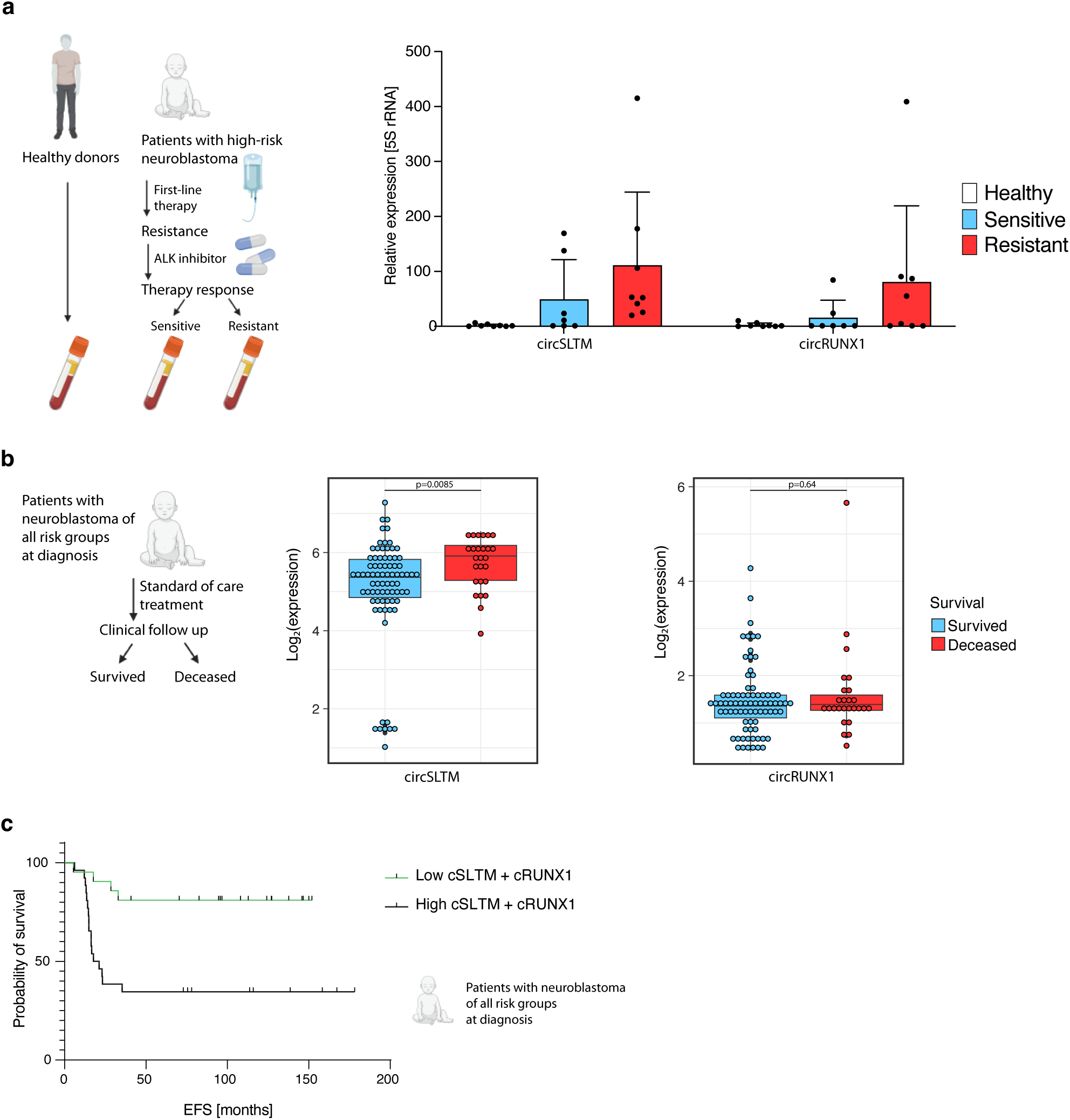
Patients with neuroblastoma and poor outcome show a higher abundance of resistance-associated circRNAs. **a,** Liquid biopsies of healthy donors (n=10 biologically independent samples) and subsequent samples of patients with high-risk neuroblastoma either sensitive (n=3 biologically independent samples) or resistant to ALKi (n=3 biologically independent samples) were analyzed by qRT-PCR. The abundance of circSLTM(3,4,5) and circRUNX1(6,7) are shown. Data are presented as mean ±SD. **b,** Tumor biopsies taken at diagnosis of 104 patients with neuroblastoma were analyzed by RNA-seq. Patients that were alive (n=78 biologically independent samples) or deceased (n=26 biologically independent samples) following their treatment_2_w_5_ere compared. Shown is the expression of circSLTM(3,4,5) and circRUNX1(6,7) in the 2 groups. **c,** The patients of b were divided in a group with both circSLTM(3,4,5) and circRUNX1(6,7) low (n=21 biologically independent samples) vs. high (n=26 biologically independent samples) expression and the probability of event-free survival (EFS) in months is shown as a Kaplan-Meier curve. In boxplots the center line represents the median, boxes indicate the interquartile range, the whiskers show the 1.5 interquartile range. Source data are provided as a source data file.

To assess the expression of the two circRNAs in an additional independent clinical cohort, we re-analyzed total RNA short-read sequencing data of 104 patients with neuroblastoma that we have previously generated [8] representing all major risk groups. Tumor biopsies were obtained at diagnosis before any given treatment. Patients were separated in 2 groups dependent on their clinical outcome in deceased patients and patients that were still alive at the end of the follow-up period. This analysis showed a higher expression of circSLTM (2,3,4) in patients with worse outcomes **(Fig. 5b)**. Further, a high expression of both circSLTM (2,3,4) and circRUNX1 (6,7) was associated with a shorter EFS **(Fig. 5c)**, as we have detected in ALK+ ALCL patients. Taken together, these data indicate that the circRNAs with higher expression in ALK+ ALCL can be detected in clinical samples of neuroblastoma and are upregulated in those patients resistant to ALKi.

## Discussion

This work highlights the use of a long-read sequencing approach to detect full-length circRNAs in a clinical cancer context for *de novo* biomarker discovery. Using preclinical models of the pediatric cancer ALK+ ALCL, circSLTM (2,3,4) and circRUNX1 (6,7) were identified to be associated with ALKi resistance. Analysis of patient samples of ALK+ ALCL revealed an association with a poor outcome and a higher abundance in liquid biopsies of patients resistant to ALKi. These results are further supported by the higher abundance of the two circRNAs in high-risk and ALKi resistant patients with the second *ALK* -related pediatric cancer neuroblastoma.

The advantage of Nanopore long-read sequencing to discover full transcript isoforms has been repeatedly demonstrated for linear transcripts but remains understudied for full-length detection of complex noncoding RNAs, such as circRNAs. Our approach allowed us to identify a high number of circRNA isoforms, with more AS events present in ALKi resistant cell models. This confirms the applicability of our sequencing protocol to detect full-length circRNAs including alternatively spliced isoforms, despite the relatively low number of circRNA reads obtained for many of the candidates. Our results are comparable to previous studies, mainly on cancer cell lines, that reported many splicing events in circRNAs, including intron retention, novel exon identification and exon skipping [18]. Among the most represented ones, 3 circRNA candidates with full intron retention were previously reported [18]. Our study enriches the reference of specific circRNA AS events that will be helpful for further research.

We experimentally validated 1 AS event of each category to confirm their presence in a circRNA. The abundance of circZNF207 (7,8,10), in which exon 9 is skipped, decreased after exonuclease treatment, which suggests that this event is also present in a cognate linear RNA expressed from the same locus. Whether the higher diversity of AS events observed in resistant samples is a general feature remains to be confirmed in further cancer entities.

To perform a robust circRNA prediction, we used two bioinformatic tools that rely on different detection strategies, since it was demonstrated that detection of lowly abundant circRNAs in long-read sequencing data is challenging due to lower sequencing depth and important differences in the sensitivity of detection tools [23]. Each tool identified the same three circRNAs which were found upregulated in ALK+ ALCL cell models resistant to ALKi treatment. CircFL-seq found more isoforms than CIRI-long, but the detailed examination of predicted circRNAs showed that most of the differences came from exon borders and the BSJ annotation, where CIRI-long seemed to better collapse. All the three upregulated circRNAs were previously reported and are referenced in major circRNA databases such as circBase [24] and circAtlas [17].

With this study, we applied the full-length circRNA sequencing approach to identify new biomarkers for the clinically challenging problem of emerging resistance to ALKi in rare pediatric cancers. Starting from ALK+ ALCL cell models, we identified circRNAs upregulated in ALKi resistant preclinical models. Recent developments highlight the importance of non-invasive liquid biopsies in pediatric cancers, like neuroblastoma, as a modality to detect relapses earlier than the current standard of care [25], which consists of imaging and clinical examination. While these studies mostly detected cell-free DNA, such as circulating *MYCN* or *ALK* [26], only few studies profiled cell-free RNA, which might be due to the technical challenge to detect generally low levels of RNA in blood. It was shown that the detection of circulating mRNAs of the known neuroblastoma markers *PHOX2B*, *TH*, *DDC*, *CHRNA3* and *GAP43* is linked to a poor outcome [27]. Further, the detection of *NPM1-ALK* fusion transcripts is clinically used as a minimal residual disease marker in ALK+ ALCL [28], but no direct association with therapy resistance was made so far. Benefiting from the increased stability of circRNAs in body fluids, we detected an increased abundance in liquid biopsies of a small cohort of ALKi resistant patients with ALK+ ALCL and also with neuroblastoma, respectively. A higher expression in tumor samples of patients with an unfavorable outcome was found for circSLTM (3,4,5) and circRUNX1 (6,7) in patients with ALK+ ALCL and neuroblastoma. A future validation of these circRNAs in independent clinical cohorts is needed to further assess their clinical utility. Moreover, the combination of currently used clinical biomarkers together with circRNAs could improve the diagnostic or prognostic accuracy in pediatric cancer diseases [29].

Recently circSLTM (3,4,5) was described in inflammatory diseases and chondrocyte function [30], suggesting a potential role in immune cell infiltration of cancers. Further, circRUNX1 (6,7) has been identified as a resistance regulator by scaffolding microRNAs (miRNA) in cancer. Specifically, circRUNX1 (6,7) was shown to regulate cisplatin and taxol drug resistance in lung cancer, which recurrently shows *ALK* aberrations [31, 32], and its expression was associated with a poor prognosis in a series of different cancer entities, including pancreatic ductal adenocarcinoma [33], but not directly with therapy resistance. Of note, the only circRNA that was so far associated with therapy resistance in pediatric cancer, is circDLGAP4, namely in neuroblastoma [9].

In conclusion, despite circRNAs are maintained at low levels in cancer and highly proliferative cells [34], our results demonstrate the feasibility of using long-read sequencing to detect full-length circRNAs and their complex splice variants in a disease context, but also the promise of using them as future biomarkers. This study further contributes to enriching the catalog of full-length circRNAs and associated AS events. Further investigation of these circRNAs in prospective clinical trials could expand our understanding of *ALK* -related pediatric cancers and improve detection of prognosis-limiting therapy resistance early.

## Methods

### Patients and biomaterial samples

This research study was conducted in accordance with the Declaration of Helsinki and Good Clinical Practice. Informed consent was obtained from the patients or their guardians. Collection and use of patient samples was approved by the institutional review boards of Institut Gustave Roussy and Charité - Universitätsmedizin Berlin within the trials and registries, respectively. Patients with ALK+ ALCL were enrolled in the European clinical trial ALCL 99 (NCT00006455) and serum samples of 4 patients were collected. 3 of the patients were boys, 1 patient was a girl. The average age was 4.25 years. Patients with neuroblastoma were registered and treated according to the German neuroblastoma clinical trials NB2004 or the German Neuroblastoma Registry (NB Registry 2016) [35]. Blood plasma samples from 7 patients with neuroblastoma were collected in the Charité Liquid Biopsy biobank (CharLi, local ethics approval: EA2/055/17) and within the Collaborative Research Center “Decoding and Targeting Neuroblastoma Evolution” CRC1588 (ethics approval: EA2/010/23). 4 of the patients were female and 3 male. The average age was 2 years. Tumor samples were staged according to the International Neuroblastoma Staging System (INSS) [36] and patient risk was defined according to the International Neuroblastoma Risk Group (INRG) [37, 38]. Gender information were not available for the patients. Patient characteristics are reported in **Supplementary Data 1**. Blood samples from 10 healthy adult donors were provided by the Etablissement fraņcais de Sang (Toulouse, France, ethical agreement number: 21PLER2021-007).

### Preparation of cell-free RNA samples

Collected plasma or serum was double-spun to obtain platelet-free plasma or serum, respectively. Briefly, peripheral blood was centrifuged at 1,900 x g for 7 minutes to separate plasma/serum from cells. All samples were centrifuged a second time at 3,250 x g for 10 minutes to remove cellular debris and platelets before RNA isolation, or storage at -80°C. Cell-free RNA (cf-RNA) was isolated using the miRNeasy Serum/Plasma Advanced Kit (Qiagen, Venlo, NL) according to the manufacturer.

### Cell lines, culture and establishment of resistant cell models

The human ALK+ ALCL cell lines SU-DHL-1, Karpas-299 and SUP-M2 were obtained from the German Collection of Microorganisms and Cell Culture GmbH (DSMZ, Braunschweig, Germany). The ALK+ ALCL COST cell line was previously established by us [39]. Cells were cultured in RPMI 1640 medium with GlutaMAX and supplemented with 20 % of heat-inactivated fetal calf serum, 1 mM sodium pyruvate and 1 % of penicillin/streptomycin (all Thermo Fisher Scientific, Waltham, MA, USA) and incubated at 37 °C, 100 % humidity and 5 % CO2.

ALKi resistant ALK+ ALCL cell models were created by treating cell lines with increasing doses of crizotinib (Selleckchem, Houston, TX, USA). Treatment started with 10 nM of crizotinib and the dose was increased only when the cells continued proliferating for several days. By this approach, SU-DHL-1 and COST cells were finally cultured in 100 nM of crizotinib and 200 nM, respectively. Resistant clones of Karpas-299 cells (cultured in 600 nM of crizotinib) and SUP-M2 (cultured in 300 nM crizotinib) were a gift from Carlo Gambacorti [40]. Together, we obtained 4 pairs of ALK+ ALCL cell models sensitive or resistant to ALKi for this study.

### Detection and validation of circRNAs and linear RNAs by PCR and Sanger sequencing

RNA from cell lines was isolated using Trizol (Thermo Fisher Scientific) and was reverse transcribed as described previously [8] with the Maxima H Minus First Strand cDNA Synthesis kit with dsDNase (Thermo Fisher Scientific) according to the manufacturer, including a DNase digest and random hexamer priming. For cell line samples, 1 µg of RNA was used as input. The cDNA was diluted 1:5 in nuclease-free water. For liquid biopsies, 8 µl of isolated cf-RNA served as input for reverse transcription, and the cDNA was diluted 1:3 with nuclease-free water.

qRT-PCR was performed using the SYBR Select Mastermix (Thermo Fisher Scientific) on a StepOnePlus real-time PCR System or a QuantStudio 5 real-time PCR System (both Thermo Fisher Scientific) following the recommendations of the manufacturer. Data were analyzed using the Design and Analysis Software (v2.4.3), or the StepOnePlus Software (v2.3, both Thermo Fisher Scientific). Divergent primers targeting the back-splice junction were designed to specifically detect circRNAs. Exon-spanning primers were designed to amplify linear RNAs. To specifically detect splicing events, primers spanning the respective exon-intron, or exon-exon junction, were designed. *MLN51* and *ABL* served as reference genes for cell line models, and 5S rRNA for liquid biopsies. The sequences of primer pairs are reported in **Supplementary Data 2**. The correct amplification of the back-splice junction of circRNAs was validated by analyzing generated amplicons by Sanger Sequencing (Eurofins Genomics GmbH, Eberberg, Germany). The circular character of circRNAs was tested by treating RNA with the exonuclease RNaseR (Lucigen, Madison, WI, USA) that digests linear RNAs followed by qRT-PCR in comparison to linear RNAs.

### Sequencing of full-length circRNAs with Oxford Nanopore

Long-read sequencing of circRNAs was performed as previously described [16] and a detailed step-by-step protocol was published (https://www.protocols.io/view/generation-of-full-length-circrna-libraries-for-ox-rm7vzy8r4lx1/v3) [41]. Briefly, circRNAs were enzymatically enriched in total RNA from cell lines and a cDNA library is created that is PCR amplified. The library serves as input for the Oxford Nanopore ligation-based sequencing protocol with native barcoding. Four cell line samples were multiplexed and sequenced on one MinION flow cell with the MinION MK1C sequencer using the MinKNOW software (v22.05.08, Oxford Nanopore, Oxford, UK). Basecalling was performed using the high accuracy model with standard parameters to create fastq files.

### Cleaning of Nanopore sequencing data

Nanopore passed fastq reads were cleaned from their adapters using Cutadapt v4.5 while considering 20 % of possible errors (-e 0.2), keeping only reads containing adapters (–discard-untrimmed parameter) and longer than 30 (-m 31). The adapters were removed by using cutadapt twice, the first time with -g AAGCAGTGGTAT-CAACGCAGAGTAC then, on the remaining reads, with -a GTACTCTGCGTTGAT-ACCACTGCTT.

### Identification of circRNAs in long-read sequencing data

CircRNAs from Nanopore long-read sequencing were detected using the CIRI-long and circFL-seq software packages. CIRI-long [15] was used as previously described [16, 41]. After cleaning reads from adapters as described above, the CIRI-long algorithm was used to search circRNA consensus sequences. To reinforce our results, we also used circFL-seq as described by Liu *et al*. [14]. We first applied the circfull RG function directly on cleaned fastq files to identify full-length circRNAs. In parallel, we applied the circfull DNSC function (*de novo* self correction method) to get consensus sequences. Second, we used circfull cRG to refine full-length circRNA prediction from consensus sequences. We finally merged the step 1 and 2 results with circfull mRG function and annotated them with circfull anno. The result is a count table of all detected different circRNA isoforms and an annotation file for each sample. In order to proceed to differential expression analysis, we merged the counts all together using the provided custom script (merge samples.sh). From the global isoform count table obtained after merging samples, we also computed a count table aggregated by circRNA. Observing that, with circFL-seq, we obtained a lot more circRNAs than with CIRI-long with very similar BSJ boundaries, so we decided to collapse the cir-cRNA predictions made by circFL-seq. The list of circFL-seq aggregated circRNAs was obtained by collapsing together very close circRNAs having both their left and right part of the BSJ at a distance of less than 10 nt.

To extract the circRNA events detected by both circFL-seq and CIRI-long, we compared the global circRNA collapsed count table of circFL-seq **(Supplementary Data 3)** with the CIRI-long count table **(Supplementary Data 4)**. As previously described for circFL-seq, we consider as identical circRNAs having their BSJ coordinates at a distance of less than 10 nt from each other. With this procedure, several circRNAs from one tool can be associated with a single circRNA from the other tool. Therefore note that this procedure generates slight differences in the number of cir-cRNAs at the intersection between the 2 tools, depending on whether we look at circFL-seq or CIRI-long. Quantitative Venn diagrams were drawn with the shiny web application of the R package eulerr (v7.0.0). For the common circRNAs at the intersection, we computed the average circRNA length and the number of different circRNAs by cell line.

The annotation of circRNAs and AS events listed in the text follows the recently proposed new nomenclature of circRNAs [42] and uses the Ensembl canonical transcriptome v108 as a reference. We further report the ID from the circAtlas 3.0 database (see below).

### Detection of circRNA hotspot regions

To check whether some specific chromosomes produce a high number of circRNAs, we computed a normalized density of circRNAs by chromosome (*Nchr c*) and for both sensitive and resistant groups, while the raw number of circRNAs detected by both tools (#*circRNAs*) was normalized by the total coding gene length by chromosome ( *codingGene length*) and by a factor of 1 million as following: *Nchr_c_* = 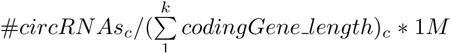, with *k* the number of coding genes by chromosome. The total coding gene length by chromosome is the sum of length (from genomic coordinates) of all genes annotated as “protein coding” into the gencode annotation file. Again for the list of common circRNAs, we calculated the number of different isoforms produced by gene and graphically represented the ones having more than 15 distincts isoforms for at least one group among sensitive and resistant cell lines. Note that we extracted the number of isoforms from circFL-seq annotations.

### Comparison of circRNAs with annotated circRNAs in circAtlas 3.0

To reveal whether the detected circRNA candidates were novel or previously annotated, we compared our results with the database circAtlas 3.0 [17] that contains more than 3.1 million annotated circRNAs across 10 species. We downloaded the human bed v3.0.zip file from circAtlas 3.0 and kept the three first fields corresponding to the chromosome number, start and end coordinates of the circRNA BSJ. A bed file containing the chromosome number, the start, and end coordinates of each circRNA BSJ was also created from the list of circRNA candidates common to circFL-seq and CIRI-long. We used the two bed files as inputs to JVenn [43] to identify circRNAs common and specific to circAtlas and our candidates. Throughout the manuscript, we mention the circRNA IDs from circAtlas 3.0.

### Detection of alternative splicing events

The CIRI-long circRNA candidates restricted to those common with circFL-seq were analyzed to identify AS events. We used bedtools [44] on both bed files of circRNA candidates and different annotation bed files generated from the canonical transcriptome file from the Ensembl genome browser (http://www.ensembl.org/). The canonical transcriptome represents in general the longest isoform of a gene, including all annotated exons. Two canonical transcriptomes were generated from the Ensembl GRCh38.108 gtf annotation file, the first one by selecting exons tagged as ”Ensembl canonical” for protein coding and non coding exons, and the second one with the additional filter of ”protein coding” for gene biotype to obtain only protein coding exons. We first used gtftools [45] to generate three bed files representing respectively only the introns of the canonical transcriptome, only the protein coding merged exons of the canonical transcriptome and finally, the union of the protein coding and non coding merged exons. Considered AS events were full intron retention, novel exons (not annotated as exon in the protein coding and non coding canonical transcriptome) and exon skipping (annotated as exon in the protein coding canonical transcriptome but not identified in the circRNA).

Retained intron events were then computed by comparing exonic circRNA candidates with introns of the canonical transcriptome. Only fully retained introns were considered as AS events. Novel exons were computed by comparing circRNA candidate exons with exons of the canonical transcriptome (protein coding and non coding exons), keeping as novel exons only the ones identified in the circRNA but not annotated in the canonical transcriptome and checking for flanking GT and AG splicing sites at their 5’ and 3’ boundaries. For each circRNA isoform, exon skipping events were computed by searching exons annotated in the canonical transcriptome (expected within the circRNA bounds) but not identified in the circRNA isoform.

### Known and novel AS events by comparison with VastDB

Identified AS events were compared with annotated events in the database VastDB [19]) to reveal whether they were novel. VastDB is a database of AS profiles across multiple tissue and cell types in different species. It includes exon skipping, novel exons and retained intron events. VastDB makes available a bed file of human AS events. We filtered the bed file of VastDB human events and generated three bed files containing, for each class of our identified AS events (retained introns, exon skipping events and new exons), the chromosome number and the start and end coordinates of each individual AS event. We also generated a bed file for each class of events detected in this study. We used bedtools to identify similar AS events by comparing the VastDB bed file to the one built from this study for the same class.

### Differential expression analysis

As we wanted to have a global overview of the isoforms diversity in our samples regarding the resistance to crizotinib, we focused our differential expression analysis on the common circRNA candidates taking the isoform counts from circFL-seq. As the DESeq2 model [46] already corrects for the number of reads matching circRNA events, we worked directly on the raw isoform counts table computed from circFL-seq after merging our samples. The R code of the analysis is provided in our github (see Code availability section). Briefly, we used the DESeq2 R package to extract the differentially expressed circRNA isoforms. We first filtered out the isoforms detected in only 1 sample (on the total of 8), which left 2,651 circRNAs candidates for the analysis. We then extracted the circRNA isoforms with a pvalue *<* 0.05.

### Illumina short-read RNA sequencing and analysis

Total RNA sequencing (RNA-seq) libraries of the 4 pairs of ALK+ ALCL cell line models (n=8) were prepared at the iGenSeq core facility of the Institut du Cerveau (Paris, France). The NEBNext rRNA depletion kit and NEBNext Ultra II Directional RNA Library kit (both New England Biolabs, Ipswich, MA, USA) were used following manufacturer’s recommendations. Libraries were pooled and sequenced on a NovaSeq 6000 S4 flow cell (Illumina, San Diego, CA, USA) with a read length of 150 bases and a sequencing depth of 2×200 million paired-end reads per sample after demultiplexing. Raw fastq files were submitted to Cutadapt (v 1.18) for low-quality trimming (-q 10,10) and excluding sequences shorter than 30 nt after trimming (-m 31).

Global gene expression was quantified using Kallisto (version 0.46.1) with the v108 Ensembl reference transcriptome (cdna+ncrna), followed by the R function tximport [47] for computing gene-level raw counts and length scaled TPM values. We finally did a log2 transformation on gene-level normalized counts with a pseudocount of 1 to compute the circRNA candidates’ host genes boxplots of expression.

To investigate the expression of circRNAs in an independent cohort of patients with neuroblastoma, we downloaded directly the circRNA count table created with CIRI2 [48] of our previously published neuroblastoma total RNA-seq dataset of 104 patients [8] and verified the association of the expression of circRNA candidates with patient outcome.

### kmer-based identification of circRNAs

To rapidly quantify our circRNA isoform candidates in an independent cohort of patients with ALK+ ALCL, we re-analyzed our previously published total RNA-seq dataset of 39 patients [20] with our recently published k-mer based quantification approach [22] via the Transipedia web interface (https://transipedia.org/) and associated circRNA expression with the histological subtype (information was available only for 38 patients). In a first step, the 38 ALCL patients corresponding RNA-seq samples were indexed using Reindeer [49] with a k-mer length of 31 nt. The index was then loaded on our personal server to have direct restricted access via Transipedia.org. In short-read RNA-seq data, circRNAs have a confident characterization only at their BSJ. We therefore designed 40-nt long k-mers centered on our candidate circRNA BSJ, to be able to quantify them in our 38 ALCL patients index. The count table is given in the source data. The count values are normalized by billion of k-mers in each sample.

### Further statistical analysis

Statistics for bioinformatics analysis is stated in the respective methods paragraph. If not stated otherwise, all *in vitro* experiments were performed in 3 independent biological replicates. Results generated by qRT-PCR were analyzed using the dCT method (when comparing to the average of control samples, for example samples of healthy donors) or the ddCT-method (when comparing to the average of untreated conditions in RNaseR treatments).

Statistical significance among or between treatment groups in *in vitro* experiments was determined by one-way (to analyze 1 parameter) or two-way ANOVA (to analyze more than 1 parameter) for more than 2 groups, or unpaired t-tests for 2 groups, using Microsoft Excel 2016 or GraphPad Prism 10 (GraphPad Software).

For survival analysis, optimal circRNA expression cutoff were determined using Cut-offFinder via the website https://molpathoheidelberg.shinyapps.io/CutoffFinder v1/ [50] using receiver operator characteristics (ROC) curve by minimizing the Manhattan distance of data points. To compute the survival curve for the intersection between our 2 circRNA candidates, we kept only the patients that belonged to the same expression group for both candidates circSLTM (3,4,5) and circRUNX1 (6,7), both above or both below their estimated optimal expression cutoff. Survival curves were computed with GraphPad Prism 10 (GraphPad Software) and compared using a log-rank (Mantel-Cox) test. For all tests, a p-value *<* 0.05 was considered as statistically significant. Graphs show the mean and error bars representing standard deviation. In box plots the center line represents the median, boxes indicate the interquartile range, the whiskers show the range. \**p <* 0.05, \*\**p <* 0.01, \*\*\**p <* 0.001, \*\*\*\**p <* 0.0001 and ns, not significant.

## Data availability

The genome reference hg38 (https://ftp.ensembl.org/pub/release-109/fasta/homo sapiens/dna/Homo sapiens.GRCh38.dna.alt.fa.gz) was downloaded from the ensembl website (https://ensemblgenomes.org) and the transcriptome annotation Gencode v41 from the gencode website (https://www.gencodegenes.org/human/release 41.html). Previously generated total RNA-seq data of patients with neuroblastoma [8] were downloaded from the European Genome-phenome Archive (EGA) under accession codes EGAS00001004022, EGAS00001005604. Previously published RNA-seq data of patients with ALK+ ALCL were downloaded from [20]. Further RNA-seq and Nanopore sequencing data generated in this study have been publicly deposited at the NCBI Gene Expression Omnibus (https://www.ncbi.nlm.nih.gov/geo) under accession numbers GSE260564 and GSE266457. The remaining data are available within the Article, Supplementary Information or Source Data file. Further information and requests for resources and reagents should be directed to and will be fulfilled by the Lead Contact, Fabienne Meggetto. Source data are provided with this paper.

## Code availability

Scripts and code used for sequencing data analysis in this study are fully referenced in the methods section. We further deposited all the code to reproduce our analysis on Github (https://github.com) under the following link: https://github.com/ Steffen-Fuchs/Nanopore ALCL circRNA.

## Supporting information

Supplementary Data S4: CIRI-long results

Supplementary Data S3: circFL-seq results

Supplementary Data S2: oligo and k-mer sequences

Supplementary Data S1: patient cohorts

## Data Availability

The genome reference hg38 (https://ftp.ensembl.org/pub/release-109/fasta/homosapiens/dna/Homo_sapiens.GRCh38.dna.alt.fa.gz) was downloaded from the ensembl website (https://ensemblgenomes.org) and the transcriptome annotation Gencode v41 from the gencode website (https://www.gencodegenes.org/human/release41.html). Previously generated total RNA-seq data of patients with neuroblastoma were downloaded from the European Genome-phenome Archive (EGA) under accession codes EGAS00001004022, EGAS00001005604. Previously published RNA-seq data of patients with ALK+ ALCL were downloaded from (Daugrois, C. et al. Cancers, 2021). Further RNA-seq and Nanopore sequencing data generated in this study have been publicly deposited at the NCBI Gene Expression Omnibus (https://www.ncbi.nlm.nih.gov/geo) under accession numbers GSE260564 and GSE266457. The remaining data are available within the Article, Supplementary Information or Source Data file. Further information and requests for resources and reagents should be directed to and will be fulfilled by the Lead Contact, Fabienne Meggetto. Source data are provided with this paper.

https://github.com/Steffen-Fuchs/Nanopore_ALCL_circRNA

## Acknowledgements

We thank the patients and their parents for granting access to clinical samples and information used in this study and all physicians involved in biosample collection. The authors are grateful to Emeline Sarot and Nathalie Saint-Laurent (Genomic and Transcriptomic facility, Technology Cluster of the Cancer Research Center of Toulouse, INSERM-UMR1037) and to Jasmin Wünschel (Charité, Berlin) for their technical assistance. We thank our hematological laboratory technicians (Charité, Berlin), Constanze Passenheim and Nadine Sachs, for their excellent technical assistance.

This work benefited from equipment and services from the iGenSeq core facility of the Institute du Cerveau (Paris, France), and genotoul bioinformatics platform Toulouse Occitanie (Bioinfo GENOTOUL, or Bio2M platform Institute of Regenerative Medicine and Biotherapies (Montpellier, France) for storage and computing resources. C.B. was supported by a fellowship from the Fondation de France. L.B. was supported by fellowships and grants from Fondation de France, Fondation l’Oréal for Women in Science, Prolific Graine de Chercheur, Association Ellye, Fondation Toulouse Cancer santé, Fondation ARC. F.M was supported by grants from Inserm, l’association Eva pour la vie, the Federation Grandir Sans Cancer, La ligue contre le cancer, Ministère de la Santé and Institut National du Cancer (INCA, PRT-K-2022-184 CircOma), la Fondation pour la recherche Médicale (EQU202403018019) and Fonds Amgen France pour la Science et l’Humain. E.A. was supported by a grant from Labex TOUCAN/Laboratoire d’excellence Toulouse Cancer and Association Cassandra. S.E.F. is a participant in the BIH-Charité Clinician Scientist Program funded by the Charité - Universitätsmedizin Berlin and the Berlin Institute of Health. S.E.F. was supported during work by a fellowship from the Deutsche Forschungsge-meinschaft (DFG, German Research Foundation, grant no. 439441203) and a grant from Ministère de la Santé and Institut National du Cancer (INCA, PRT-K-2022-184 CircOma). This work was supported by the Deutsche Forschungsgemeinschaft (German Research Foundation) through the Collaborative Research Center “Decoding and Targeting Neuroblastoma Evolution” CRC1588 (project no. 493872418) to A.E., A.S., M.L., H.E.D., J.M.R. and S.E.F.; by the European Union (EU) and the German Federal Ministry of Research, Technology and Space (BMFTR) through the TRANSCAN-3 EXPLORE-NB consortium (01KT2401A) to H.E.D.; and by the Charité-BIH Advanced Clinician Scientist Program to H.E.D.; and by the Charité Medical Scientist Program (MSP) and the Lydia Rabinowitsch grant to A.S. Schemes were created with Biorender (www.biorender.com).

## Author contributions

L.B., E.A., J.M.R., A.E., S.P., F.M. contributed to the study design. C.B., C.G., S.E.F. collected and interpreted data. C.B., C.G., S.E.F. generated sequencing libraries and performed RNA-seq and Nanopore-seq data analysis. S.E.F. performed experiments and analyzed data. A.S., M.L., H.E.D., C.R., C.Q., V.V., L.L. provided patient samples. C.B., C.G., S.E.F. led the study design, performed the data analysis, and wrote the manuscript. All authors contributed to manuscript drafting.

