## Supplementary figures for "Identification of full-length circular RNAs linked with therapy resistance of pediatric cancers"

### Long-read sequencing identifies full-length circular RNAs linked with therapy resistance of pediatric cancers

**Figure S1| Sensitive and resistant cell models do not show differences in sequencing quality, while more circRNAs are detected in resistant models**

**a, b,** RNA of cell models from the ALKi sensitive (left panel) and resistant group (right panel) was treated enzymatically to enrich for circRNAs and deplete other RNAs. Expression levels were detected by qRT-PCR and enrichments are shown relative to the control samples (n=4 biologically independent samples each, data are presented as mean ± SD).

**c,** Number of reads in the 2 groups of cell models (n=4 biologically independent samples each). Data are represented as boxplot (blue: sensitive group; red: resistant group).

**d,** Distribution of read length in the different samples (n=8 biologically independent samples). Data are represented as boxplot (blue: sensitive group; red: resistant group).

**e,f,** Number of circRNAs detected by CIRI-Long and circFL in ALKi resistant cell models (n=4 biologically independent samples) and sensitive models (n=4 biologically independent samples).

In boxplots the center line represents the median, boxes indicate the interquartile range, the whiskers show the 1.5 interquartile range. Source data are provided as a source data file.

**Figure S2| Novel exons are more prominent in resistant cell models**

**a,** The upset plot shows the total number of novel exon events identified by Nanopore sequencing per group of cell models (blue, ALKi sensitive and red, ALKi resistant, n=4 biologically independent samples each) and the events specific or common (indicated by a connecting line) to the different cell models.

**b,** The genome browser view shows circARNTL2(2,3,4,NE,5) with a novel exon (NE) between exon 4 and exon 5. The event was validated by Sanger sequencing of the junction in Karpas-299 cells. Details on the circRNA ID (circATLAS 3.0), length and coordinates are given. Source data are provided as a source data file.

**Figure S3| Exon skipping events have a higher frequency in resistant models**

**a,** The upset plot shows the total number of exon skipping events by group of cell models (blue, ALKi sensitive and red, ALKi resistant, n=4 biologically independent samples each) and the events specific or common (indicated by a connecting line) to the different cell models. Events were identified by Nanopore sequencing.

**b,** The genome browser view shows circZNF207(7,8,10) with skipped exon 9. The event was validated by Sanger sequencing of the junction between exon 10 and 8 in Karpas-299 cells. Details on the circRNA ID (circATLAS 3.0), length and coordinates are given.

**c,** Validation of the circular character of RNAs where the splicing event was detected, was performed by treatment with RNaseR of Karpas-299 cells and subsequent qRT-PCR in comparison to linear controls (n=3 biologically independent experiments. Data are presented as mean ± SD, Two-way ANOVA test). Stable events are marked in bold. Source data are provided as a source data file.

**Figure S4| Resistant models show a higher expression of circRUNX1 and circRAD23B**

**a**, The genome browser view shows circRUNX1(6,7) and circRAD23B(3,4,5) with read coverage in ALKi sensitive (blue) and ALKi resistant (red) groups of cell models (n=4 biologically independent samples each).

**b**, **c,** Validation of circRUNX1(6,7) and circRAD23B(3,4,5) by Sanger sequencing in Karpas-299 cells. Details on the circRNA ID (circATLAS 3.0), length and coordinates are given. Source data is provided as a source data file.

**d,** The expression of cognate linear RNAs was quantified in short-read sequencing data of the same sensitive and resistant cell models as in **a.**

In boxplots the center line represents the median, boxes indicate the interquartile range, the whiskers show the 1.5 interquartile range. Data are represented as a boxplot. In boxplots the center line represents the median, boxes indicate the interquartile range, the whiskers show the 1.5 interquartile range. Source data are provided as a source data file.

Supp Fig. 1

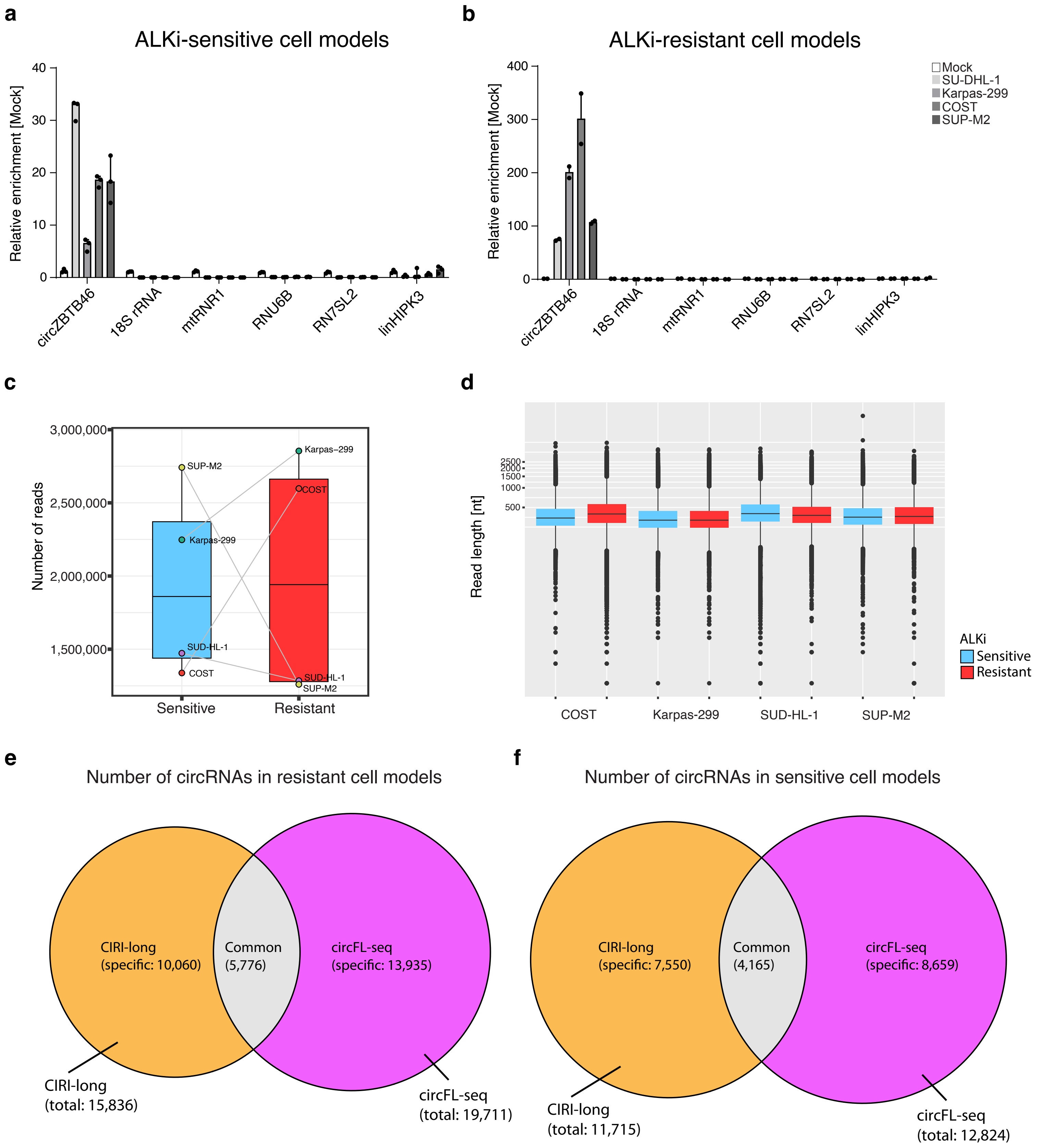

a
ALKi-sensitive cell models
Relative enrichment [Mock]
circZBTB46
18S rRNA
mtRNR1
RNU6B
RN7SL2
linHIPK3
b
ALKi-resistant cell models
Mock
SU-DHL-1
Karpas-299
COST
SUP-M2
c
Number of reads
SUP-M2
Karpas-299
SUD-HL-1
COST
Sensitive
Resistant
d
Read length [nt]
ALKi
Sensitive
Resistant
COST
Karpas-299
SUD-HL-1
SUP-M2
e
Number of circRNAs in resistant cell models
CIRI-long (specific: 10,060)
Common (5,776)
circFL-seq (specific: 13,935)
CIRI-long (total: 15,836)
circFL-seq (total: 19,711)
f
Number of circRNAs in sensitive cell models
CIRI-long (specific: 7,550)
Common (4,165)
circFL-seq (specific: 8,659)
CIRI-long (total: 11,715)
circFL-seq (total: 12,824)

Supp Fig. 2

**a**

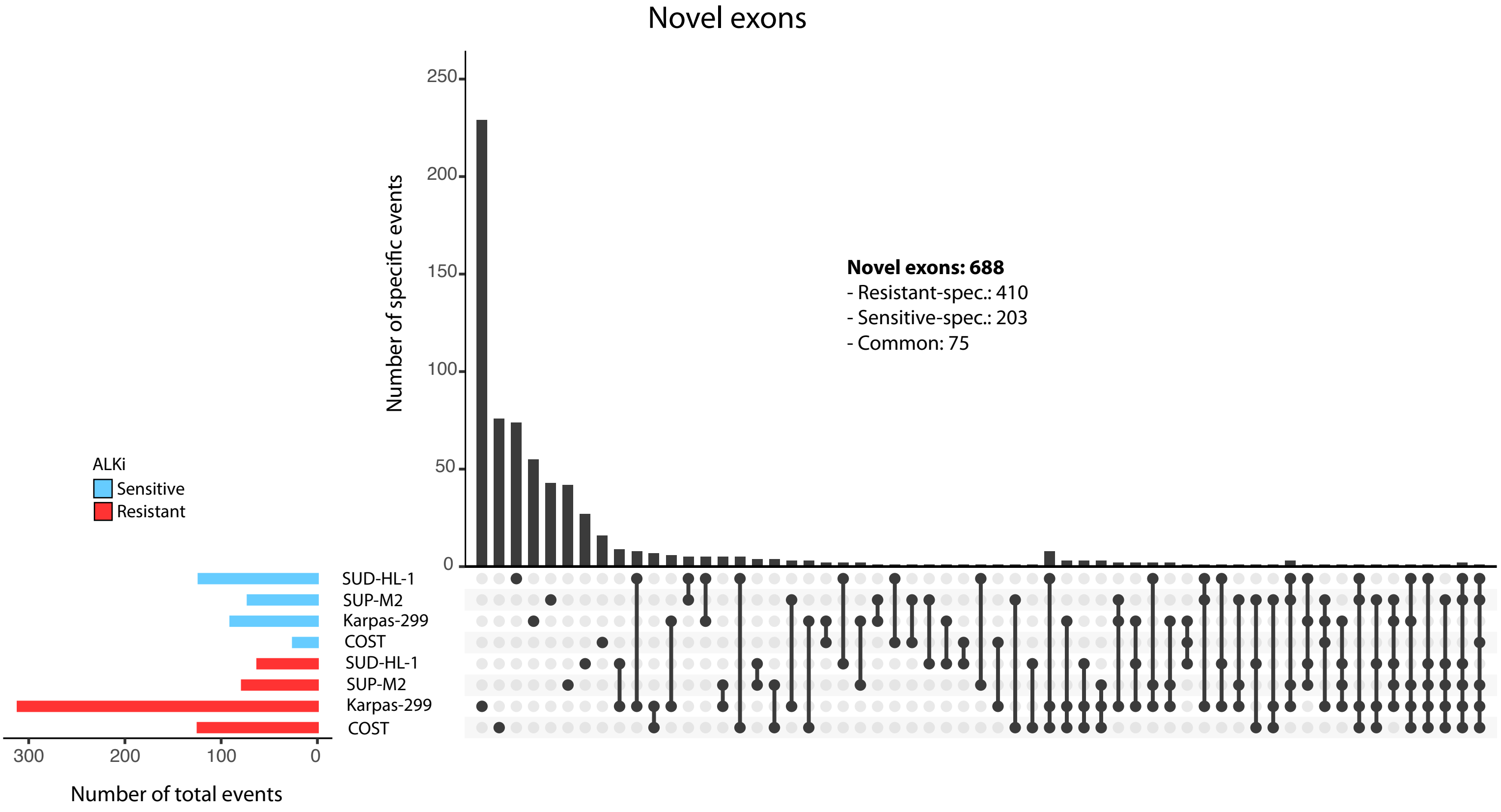
Novel exons
Number of specific events
250
200
150
100
50
0
Novel exons: 688
- Resistant-spec.: 410
- Sensitive-spec.: 203
- Common: 75
ALKi
Sensitive
Resistant
SUD-HL-1
SUP-M2
Karpas-299
COST
SUD-HL-1
SUP-M2
Karpas-299
COST
300
200
100
0
Number of total events

**b**

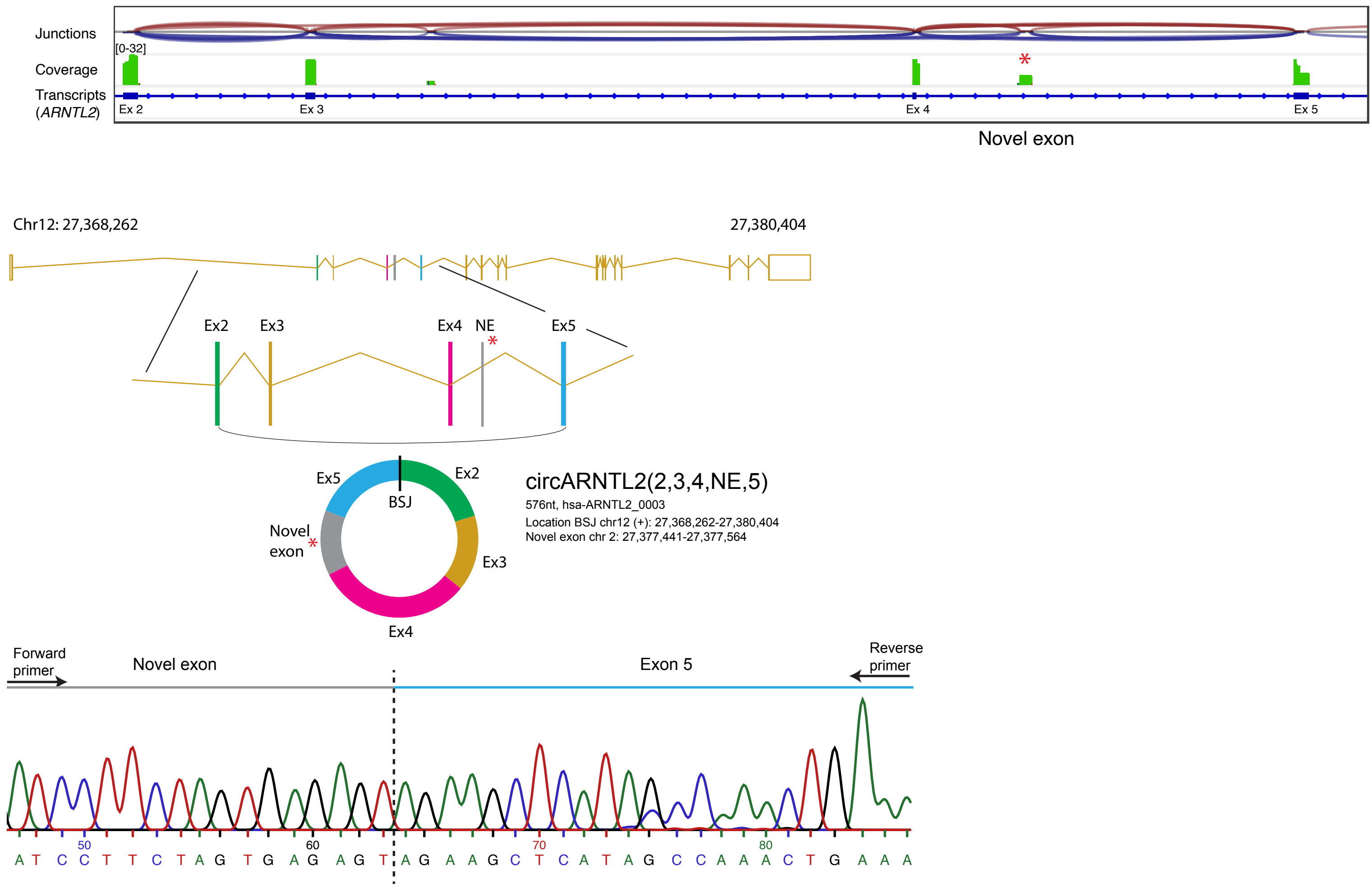
Junctions
[0-32]
Coverage
Transcripts
(ARNTL2)
Ex 2
Ex 3
Ex 4
Ex 5
*
Novel exon
Chr12: 27,368,262
27,380,404
Ex2
Ex3
Ex4
NE
Ex5
Ex5
Ex2
BSJ
Novel exon
Ex3
Ex4
circARNTL2(2,3,4,NE,5)
576nt, hsa-ARNTL2_0003
Location BSJ chr12 (+): 27,368,262-27,380,404
Novel exon chr 2: 27,377,441-27,377,564
Forward primer
Novel exon
Exon 5
Reverse primer
50
60
70
80
A T C C T T C T A G T G A G A G T A G A A G C T C A T A G C C A A A C T G A A A

Supp Fig. 3

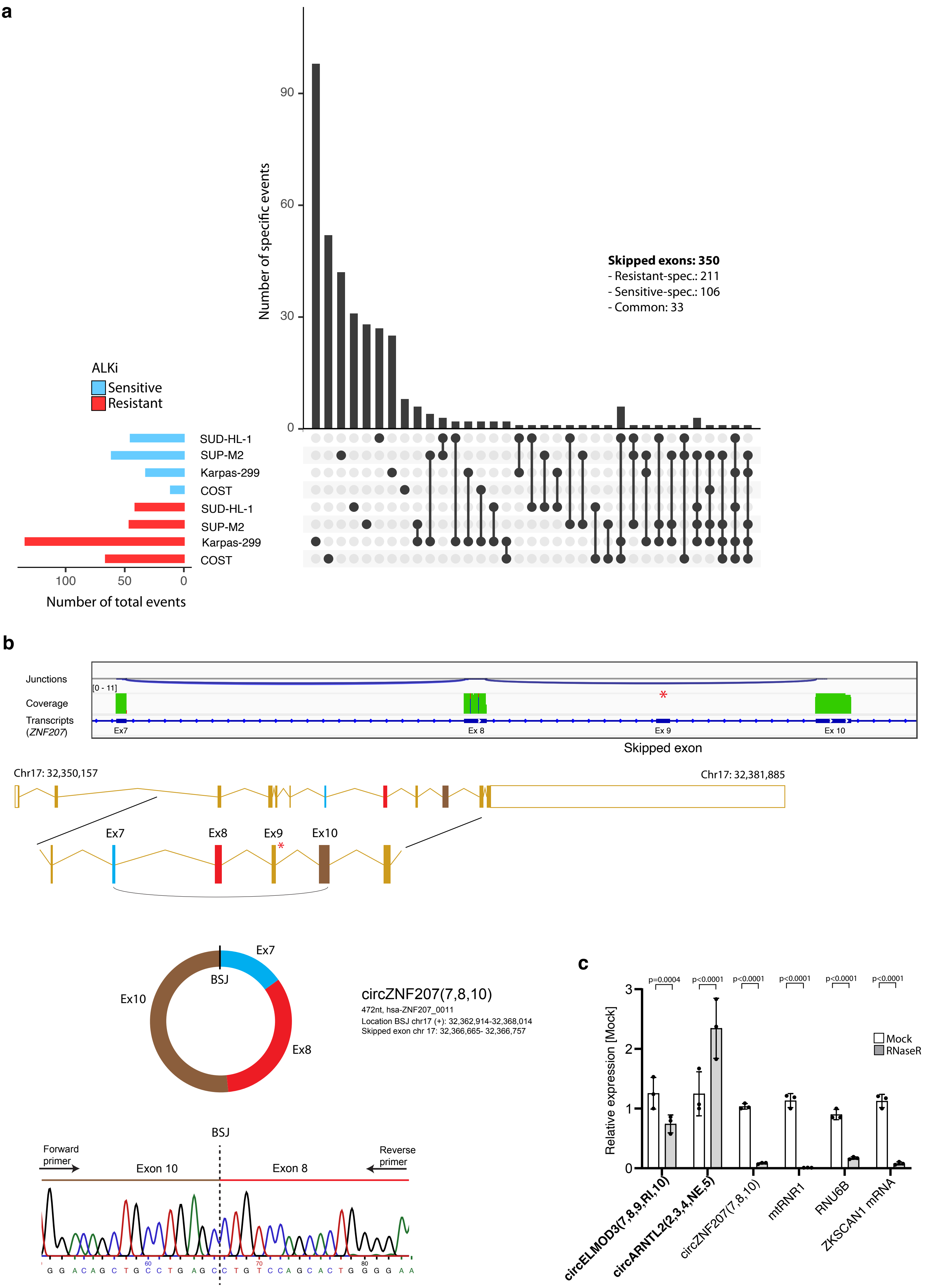

a
Exon skipping
Number of specific events
90
60
30
0
Skipped exons: 350
- Resistant-spec.: 211
- Sensitive-spec.: 106
- Common: 33
ALKi
Sensitive
Resistant
SUD-HL-1
SUP-M2
Karpas-299
COST
SUD-HL-1
SUP-M2
Karpas-299
COST
100
50
0
Number of total events
b
Junctions
[0 - 11]
Coverage
Transcripts
(ZNF207)
Ex7
Ex 8
Ex 9
Ex 10
Skipped exon
Chr17: 32,350,157
Chr17: 32,381,885
Ex7
Ex8
Ex9
Ex10
Ex7
BSJ
Ex10
Ex8
circZNF207(7,8,10)
472nt, hsa-ZNF207_0011
Location BSJ chr17 (+): 32,362,914-32,368,014
Skipped exon chr 17: 32,366,665- 32,366,757
BSJ
Forward primer
Exon 10
Exon 8
Reverse primer
c
p=0.0004
p<0.0001
p<0.0001
p<0.0001
p<0.0001
p<0.0001
Relative expression [Mock]
3
2
1
0
Mock
RNaseR
circELMOD3(7,8,9,RI,10)
circARNTL2(2,3,4,NE,5)
circZNF207(7,8,10)
mtRNR1
RNU6B
ZKSCAN1 mRNA

Supp Fig. 4

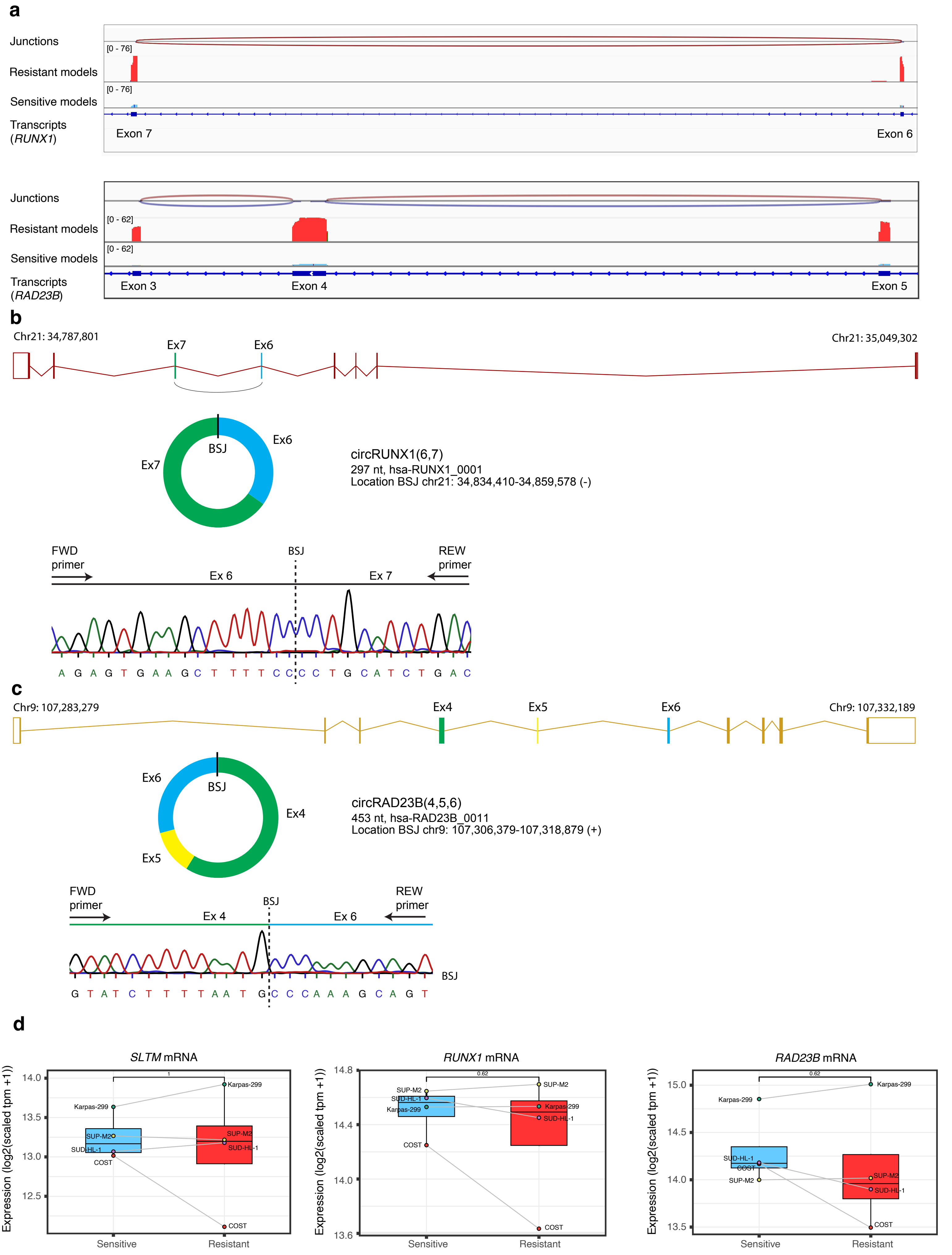
a
Junctions
Resistant models
Sensitive models
Transcripts (RUNX1)
[0 - 76]
[0 - 76]
Exon 7
Exon 6
Junctions
Resistant models
Sensitive models
Transcripts (RAD23B)
[0 - 62]
[0 - 62]
Exon 3
Exon 4
Exon 5
b
Chr21: 34,787,801
Chr21: 35,049,302
Ex7
Ex6
BSJ
Ex6
Ex7
circRUNX1(6,7)
297 nt, hsa-RUNX1_0001
Location BSJ chr21: 34,834,410-34,859,578 (-)
FWD primer
BSJ
REW primer
Ex 6
Ex 7
A G A G T G A A G C T T T T C C C C T G C A T C T G A C
c
Chr9: 107,283,279
Ex4
Ex5
Ex6
Chr9: 107,332,189
Ex6
BSJ
Ex4
Ex5
circRAD23B(4,5,6)
453 nt, hsa-RAD23B_0011
Location BSJ chr9: 107,306,379-107,318,879 (+)
FWD primer
BSJ
REW primer
Ex 4
Ex 6
BSJ
G T A T C T T T T A A T G C C C A A A G C A G T
d
SLTM mRNA
RUNX1 mRNA
RAD23B mRNA
Expression (log2(scaled tpm +1))
Sensitive
Resistant
1
0.62
0.62
Karpas-299
SUP-M2
SUD-HL-1
COST
